# Epigenetic contributions to clinical risk prediction of cardiovascular disease

**DOI:** 10.1101/2022.10.21.22281355

**Authors:** Aleksandra D Chybowska, Danni A Gadd, Yipeng Cheng, Elena Bernabeu, Archie Campbell, Rosie M Walker, Andrew M McIntosh, Nicola Wrobel, Lee Murphy, Paul Welsh, Naveed Sattar, Jackie F Price, Daniel L McCartney, Kathryn L Evans, Riccardo E Marioni

## Abstract

**Background and Aims:** Cardiovascular disease (CVD) is among the leading causes of death worldwide. Discovery of new omics biomarkers could help to improve risk stratification algorithms and expand our understanding of molecular pathways contributing to the disease. Here, ASSIGN – a cardiovascular risk prediction tool recommended for use in Scotland – was examined in tandem with epigenetic and proteomic features in risk prediction models in ≥12,657 participants from the Generation Scotland cohort.

**Methods:** Previously generated DNA methylation-derived epigenetic scores (EpiScores) for 109 protein levels were considered, in addition to both measured levels and an EpiScore for cardiac troponin I (cTnI). The associations between individual protein EpiScores and the CVD risk were examined using Cox regression (n_cases_ý1,274; n_controls_ý11,383) and visualised in a tailored R application. Splitting the cohort into independent training (n=6,880) and test (n=3,659) subsets, a composite CVD EpiScore was then developed.

**Results:** Sixty-five protein EpiScores were associated with incident CVD independently of ASSIGN and the measured concentration of cTnI (P<0.05), over a follow up of up to 16 years of electronic health record linkage. The most significant EpiScores were for proteins involved in metabolic, immune response and tissue development/regeneration pathways. A composite CVD EpiScore (based on 45 protein EpiScores) was a significant predictor of CVD risk independent of ASSIGN and the concentration of cTnI (Hazard Ratio HR=1.32, P=3.7×10^-3^, 0.3% increase in C-statistic).

**Conclusions:** EpiScores for circulating protein levels are associated with CVD risk independent of traditional risk factors and may increase our understanding of the aetiology of the disease.

**Graphical abstract:** 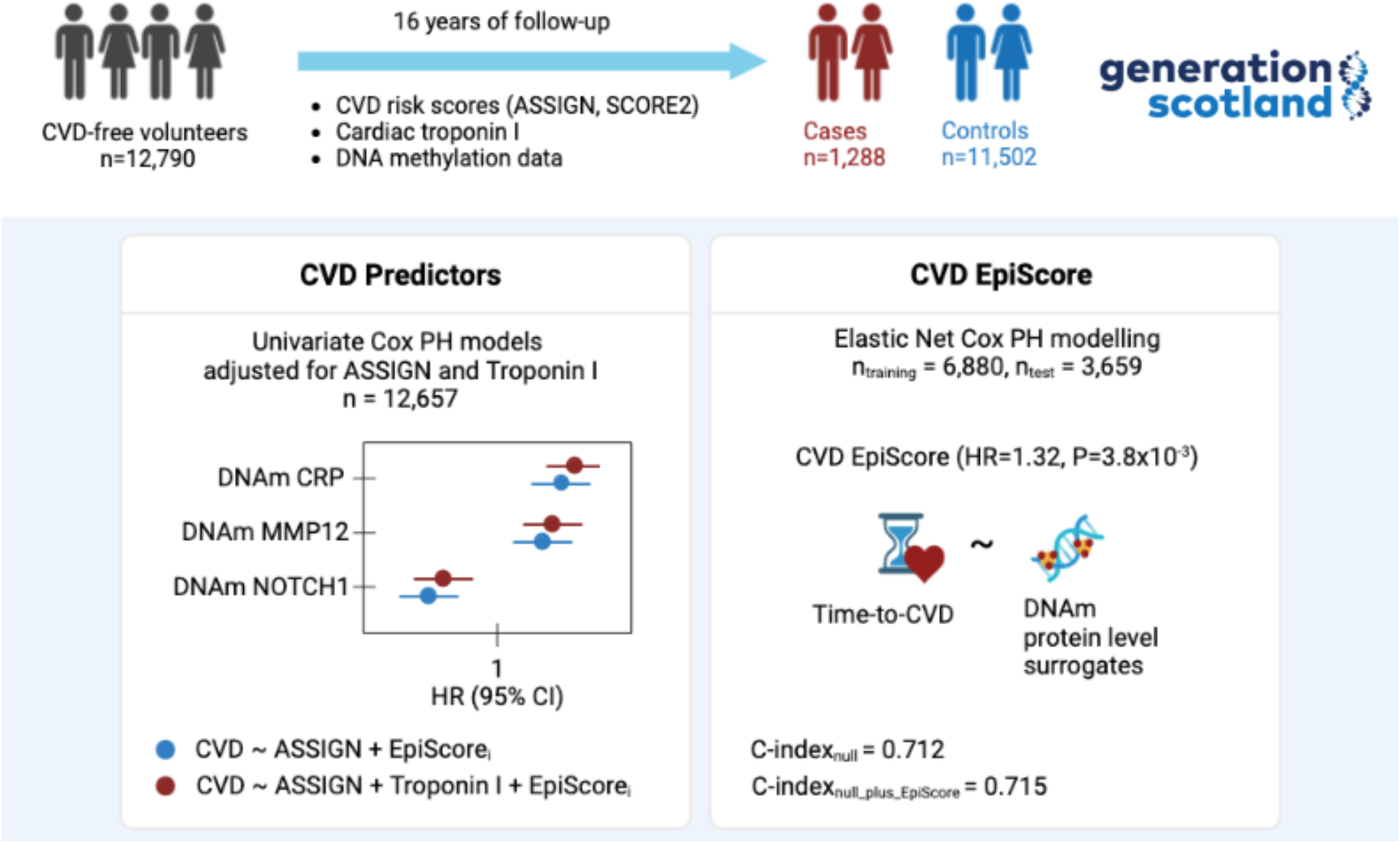

**Structural graphical abstract:** ASSIGN – a cardiovascular risk prediction tool recommended for use in Scotland – was examined in tandem with epigenetic and proteomic features in risk prediction models in ≥12,657 participants from the Generation Scotland cohort. Cox regression was used to model the association between individual predictors and CVD hospitalisation events ascertained over 16 years of follow-up. Finally, a composite protein EpiScore was developed (based on the protein EpiScores) and its predictive performance was tested. CVD – Cardiovascular Disease, EpiScore – Epigenetic Score, Cox PH – Cox Proportional Hazards Regression, DNAm – DNA methylation.

## Introduction

For the past 20 years, cardiovascular disease (CVD) has been among the leading causes of mortality and morbidity worldwide. Given that many CVD cases are preventable, it is important to identify at-risk individuals early, when an intervention is most likely to be effective, and translate this knowledge into preventative strategies ^1, 2^. Although there are many CVD risk prediction algorithms, currently they have a limited predictive performance. It may be possible to improve on that by discovering novel factors strongly associated with the disease, for example the type and the concentrations of proteins expressed as a response to the damage of the cardiovascular system.

An established and a highly sensitive marker of myocardial damage is cardiac troponin ^3^. It is a complex of three proteins, namely cardiac troponin I (cTnI), cardiac troponin T (cTnT), and cardiac troponin C (cTnC) regulating the contraction of the cardiac muscle. Cardiac forms of troponin T^4, 5^ and I are expressed almost exclusively in the heart ^6^. Following myocyte damage, cardiac troponin enters the circulation and can be detected in blood samples. A high-sensitivity cardiac troponin test plays a role in a rapid diagnosis of myocardial infarction^3^. Low-grade elevations in cardiac troponin are associated with increased risk of CVD^3^.

Individual differences in protein concentration can be well-captured by DNA methylation (DNAm). DNAm is a type of epigenetic modification characterised by the addition of methyl groups to DNA. Typically, the methyl group is added at cytosine-phosphate-guanine (CpG) dinucleotides, which are found mostly (but not exclusively) in gene promoters ^7^. Blocking promoters, to which activating transcription factors should bind in order to initiate transcription, is one of the mechanisms by which DNA methylation can precisely regulate gene expression ^8^.

DNAm-based proxies for protein levels are referred to as protein EpiScores and are broadly analogous to polygenic risk scores. These methylation scores can be derived from penalised linear regression models of protein concentrations. Due to their temporal stability, protein EpiScores may exhibit stronger associations with disease outcomes than singular protein measurements, which are known to fluctuate between measurements ^9–12^. We have shown that EpiScores for 109 circulating protein levels are associated with the time-to-diagnosis for a host of leading causes of morbidity and mortality, including cardiovascular outcomes ^13^. Protein EpiScores are therefore useful biomarker tools for disease risk stratification.

Here, we examine whether protein EpiScores, calculated for ≥12,657 participants of the Generation Scotland (GS), study can augment predictions made by a CVD risk calculator developed for use in Scotland (ASSIGN^14^). We first run individual Cox proportional hazards (PH) models to discover relationships between individual protein EpiScores and incident CVD. We then create a CVD EpiScore (based on the protein EpiScores) and test the additional predictive performance offered by it for CVD risk stratification.

## Methods

### Generation Scotland (GS)

Generation Scotland is a population-based and family-structured cohort study of individuals from Scotland ^15^. Between 2006 and 2010, patients at collaborating general medical practices in Scotland aged 35-65 years were invited to join the study. Subsequently, participants were asked to identify first-degree relatives aged 18 and over who were then invited to participate. 24,088 participants, aged 18-99 years, completed a health survey. Clinical and physical characteristics of 21,521 individuals who attended a clinic were measured using a standardized protocol. Fasting blood samples were obtained in clinic using a standard operating procedure.

### Measurement of high-sensitivity troponin

The concentrations of high sensitivity cTnI (Abbott Diagnostics) were measured in 19,130 GS individuals. Before the assay, samples were spun for 5 minutes at 2000g. The measurements were taken using i1000SR and Cobas e411 devices using the manufacturers’ quality controls and calibrators. The limit of detection set by manufacturers of these devices is 1.2 ng/L. Anything below this limit is reported as a blank value. Correcting for blank values consisted of reporting results below the threshold of blank as 0.6 ng/L ^3^.

### DNA Methylation

DNAm was profiled in blood samples using the Illumina EPIC array. Quality control details have been described previously ^16^. Filtering for poorly detected probes and samples, non-blood (e.g., saliva) samples and outliers was performed. Subsequently, non-CpG probes as well as probes on the X and Y chromosomes were removed. Missing CpG values were mean-imputed. To ensure any signatures generalise to as many cohorts as possible, the sites were subset to those also present on the older Illumina 450k array (n=453,093 CpGs). The full quality-controlled dataset contained 18,413 individuals.

Methylation profiling was carried out in three sets. Set 1 contained 5,087 individuals, Set 2 contained 4,450 individuals and Set 3 contained 8,876 individuals. Participants in Set 2 were genetically unrelated to each other and to those in Set 1 (genetic relationship matrix (GRM) threshold <0.05) while more complex relationship structures were present within and between Sets 1 and 3.

The 109 protein EpiScores described by Gadd *et al*. ^13^ were projected into the cohort via the publicly available MethylDetectR Shiny App ^17^. A short tutorial on MethylDetectR is available at https://youtu.be/65Y2Rv-4tPU. Prior to calculating the scores, methylation level at each site was scaled within set to have a mean of zero and standard deviation (SD) of one.

### ASSIGN and SCORE2 scores

ASSIGN scores were calculated for 16,366 GS individuals with complete information about the component parts (age, sex, smoking status, systolic blood pressure, total cholesterol, high density lipoprotein cholesterol, family history of premature CVD, diagnosis of rheumatoid arthritis, diagnosis of diabetes, and a deprivation score). Variables were adjusted as per official guidance (*FAQs - ASSIGN Score*, 2022). This included modifying the number of cigarettes smoked per day in recent quitters, adding 20mmHg to systolic blood pressure in individuals on blood pressure medications, and adding 10 to the number of cigarettes smoked per day in rheumatoid arthritis patients. The ASSIGN score was calculated in R using publicly available coefficients and formulae ^19^. A subset of the obtained scores was validated against the scores produced by the online tool ^14^.

SCORE2 values were calculated for 16,934 GS individuals using external coefficients ^20^. The estimates were calibrated according to region-specific scaling factors and validated against the online tool ^21^. The R scripts used to derive ASSIGN and SCORE2 are available at https://github.com/aleksandra-chybowska/troponin_episcores.

### Cardiovascular disease events

CVD cases were ascertained through data linkage to NHS Scotland hospital records. Participants were followed to the end of September 2021. A composite CVD outcome was defined as per Welsh *et al.* ^3^ and included the following *International Classification of Diseases, 10th Revision codes*: I20–25, G45, I60–69, I00–I99, L29.5, L31.1, K40–46, K49, K75. A total of 2,265 incident CVD cases were observed over a follow up period of up to 16 years **(Supplementary Figure 1).**

### Statistical analysis for risk associations

Cox proportional hazards (PH) models were used to investigate the relationship between the CVD outcome and the following potential biomarkers: measured cardiac troponin, troponin EpiScores, and 109 protein EpiScores. All models were adjusted for ASSIGN. Protein EpiScore models were additionally adjusted for the corrected concentration of cTnI (see **Methods - Measurement of high-sensitivity troponin)**. The levels of protein EpiScores as well as troponin concentrations were rank-based inverse normalised prior to the analyses. CVD cases comprised individuals diagnosed after baseline who subsequently died as well as of those who remained alive after receiving a diagnosis. Controls were censored at the end of the follow up period (September 2021) or at time of death (CVD-free survival). Models based on data from merged Sets 1, 2, and 3 were generated using coxme library (v 2.2.16) with a kinship matrix fitted as a random effect to adjust for relatedness. Cox model assumptions were checked by running the cox.zph function from the survival library (v. 3.2.7) ^22^ was used to examine Schoenfeld residuals (local and global tests with significance threshold P< 0.05). These models were not adjusted for relatedness. Survival, forest, and hazard-over-time plots were generated using a custom shiny application available at https://shiny.igc.ed.ac.uk/3d2c8245001b4e67875ddf2ee3fcbad2/.

### Composite CVD EpiScore

An epigenetics-based CVD score referred to as the CVD EpiScore was generated. It considered the 109 DNAm scores reflecting the concentrations of plasma proteins (protein EpiScores) from Gadd *et al* ^13^ as potentially informative features (n_training_=6,880, n_test_=3,659, **Supplementary Figure 2**).The score was derived using two different modelling techniques: Cox PH Elastic Net and Random Survival Forest. While Elastic Net models were trained using glmnet (v 4.0.2) ^23^, Random Survival Forest models were trained using randomForestSRC (v 3.2.0). Protein EpiScores were rank-based inverse normalised prior to training.

Elastic Net models were trained with the L1-L2 mixing parameter set to alpha=0.5 and with ten-fold cross-validation. The features with non-zero coefficients were used to generate composite score. Random Forest models were trained using ten-fold cross-validation. The 10-year onset probabilities were calculated and used as a composite CVD EpiScore.

Finally, to test whether the CVD EpiScore improved prediction of CVD over and above ASSIGN, the following Cox PH models were fit to the test set: a null model adjusted for age, sex, and ASSIGN, then full models, which included age, sex, ASSIGN score and 1) the CVD EpiScore, 2) the concentration of rank-based inverse normalised cTnI and 3) the CVD EpiScore + the concentration of cTnI.

#### Evaluating Performance of the Composite Scores

Cox PH analysis can be used model the risk of incident CVD for different prediction periods. As the ASSIGN score estimates the risk of developing CVD over 10 years, we used 10-year CVD incidence to evaluate the classification performance of our models. To calculate the binary CVD outcome, time-to-event was truncated at 10-years. Prediction probabilities were obtained in the test set for each model by calculating the cumulative baseline hazard using the Breslow estimator ^24^. Prediction metrics (Area Under the receiver operator characteristic Curve (AUC), Precision-Recall AUC and C-index) were then generated using the pROC package ^25^.

## Results

### Clinical risk prediction tools

ASSIGN scores were calculated for 16,366 individuals with non-missing risk factor data. To meet the proportional hazards assumption of the Cox model, the dataset was filtered to individuals between 30 and 70 years old (results split by decade are presented in **Supplementary Table 1)** and trimmed of outliers (points beyond 3 SDs of the mean, n=181). This left a cohort of 12,790 individuals, which was further filtered to records with non-missing concentrations of cTnI (n = 12,657). **Table 1** summarises the training, test, and full datasets.

**Table 1.**
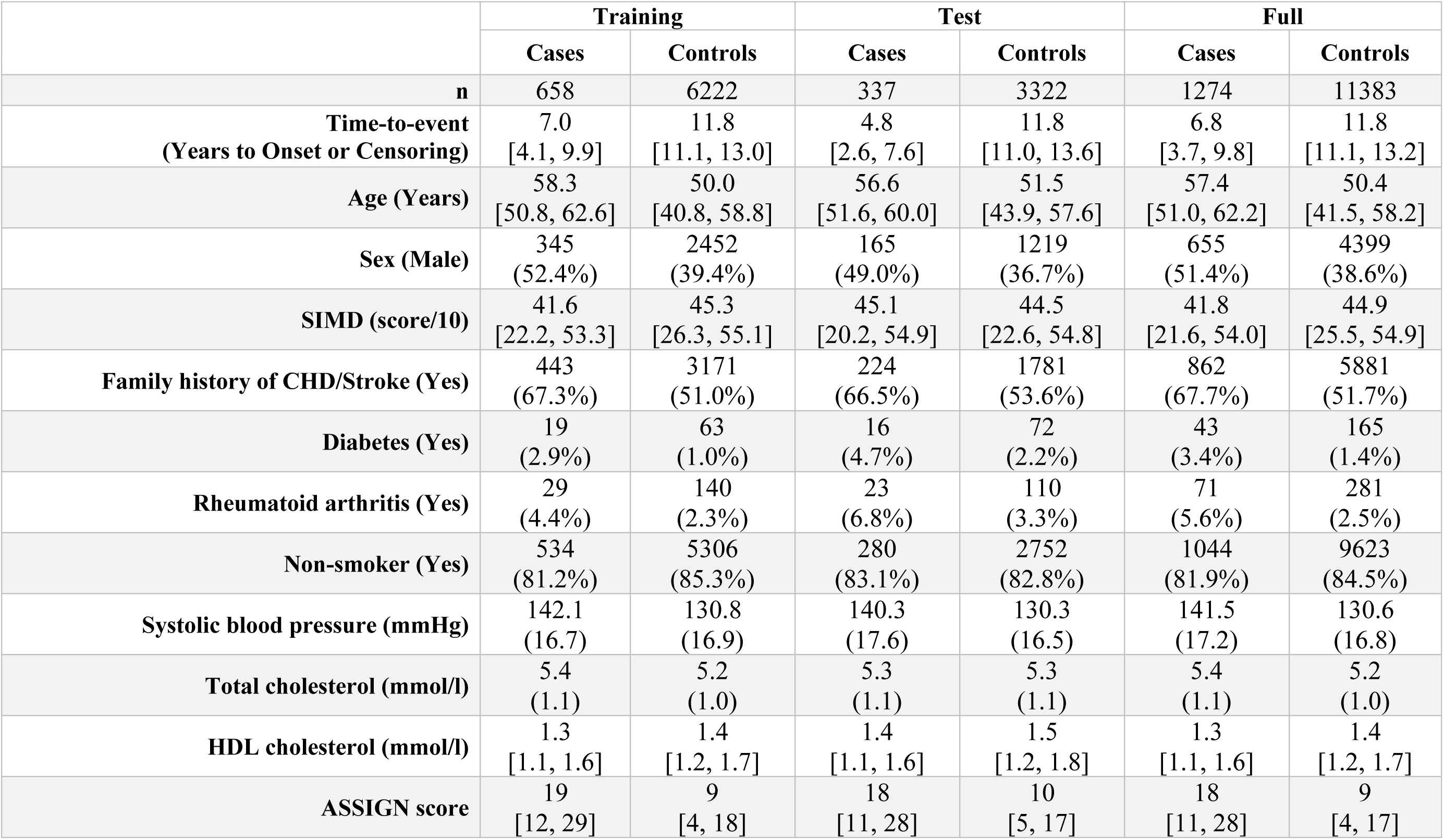
Summary of training, test, and full datasets. For continuous variables with normal distributions, summary values are reported as mean (SD). Median [Q1, Q3] are given for continuous variables which do not follow a normal distribution. A number and a percentage of samples are reported for categorical variables. SIMD **–** Scottish Index of Multiple Deprivation, CHD – Coronary Heart Disease.

|  | Training |  | Test |  | Full |  |
| --- | --- | --- | --- | --- | --- | --- |
|  | Cases | Controls | Cases | Controls | Cases | Controls |
| **n** | 658 | 6222 | 337 | 3322 | 1274 | 11383 |
| **Time-to-event**<br>**(Years to Onset or Censoring)** | 7.0<br>[4.1, 9.9] | 11.8<br>[11.1, 13.0] | 4.8<br>[2.6, 7.6] | 11.8<br>[11.0, 13.6] | 6.8<br>[3.7, 9.8] | 11.8<br>[11.1, 13.2] |
| **Age (Years)** | 58.3<br>[50.8, 62.6] | 50.0<br>[40.8, 58.8] | 56.6<br>[51.6, 60.0] | 51.5<br>[43.9, 57.6] | 57.4<br>[51.0, 62.2] | 50.4<br>[41.5, 58.2] |
| **Sex (Male)** | 345<br>(52.4%) | 2452<br>(39.4%) | 165<br>(49.0%) | 1219<br>(36.7%) | 655<br>(51.4%) | 4399<br>(38.6%) |
| **SIMD (score/10)** | 41.6<br>[22.2, 53.3] | 45.3<br>[26.3, 55.1] | 45.1<br>[20.2, 54.9] | 44.5<br>[22.6, 54.8] | 41.8<br>[21.6, 54.0] | 44.9<br>[25.5, 54.9] |
| **Family history of CHD/Stroke (Yes)** | 443<br>(67.3%) | 3171<br>(51.0%) | 224<br>(66.5%) | 1781<br>(53.6%) | 862<br>(67.7%) | 5881<br>(51.7%) |
| **Diabetes (Yes)** | 19<br>(2.9%) | 63<br>(1.0%) | 16<br>(4.7%) | 72<br>(2.2%) | 43<br>(3.4%) | 165<br>(1.4%) |
| **Rheumatoid arthritis (Yes)** | 29<br>(4.4%) | 140<br>(2.3%) | 23<br>(6.8%) | 110<br>(3.3%) | 71<br>(5.6%) | 281<br>(2.5%) |
| **Non-smoker (Yes)** | 534<br>(81.2%) | 5306<br>(85.3%) | 280<br>(83.1%) | 2752<br>(82.8%) | 1044<br>(81.9%) | 9623<br>(84.5%) |
| **Systolic blood pressure (mmHg)** | 142.1<br>(16.7) | 130.8<br>(16.9) | 140.3<br>(17.6) | 130.3<br>(16.5) | 141.5<br>(17.2) | 130.6<br>(16.8) |
| **Total cholesterol (mmol/l)** | 5.4<br>(1.1) | 5.2<br>(1.0) | 5.3<br>(1.1) | 5.3<br>(1.1) | 5.4<br>(1.1) | 5.2<br>(1.0) |
| **HDL cholesterol (mmol/l)** | 1.3<br>[1.1, 1.6] | 1.4<br>[1.2, 1.7] | 1.4<br>[1.1, 1.6] | 1.5<br>[1.2, 1.8] | 1.3<br>[1.1, 1.6] | 1.4<br>[1.2, 1.7] |
| **ASSIGN score** | 19<br>[12, 29] | 9<br>[4, 18] | 18<br>[11, 28] | 10<br>[5, 17] | 18<br>[11, 28] | 9<br>[4, 17] |

We observed that ASSIGN risk scores were strongly positively correlated with estimates produced by SCORE2 – an algorithm validated to predict 10-year CVD risk in European populations. In a sample of GS individuals within the recommended SCORE2 age range (40-69 years old, n=11,348), the Spearman’s correlation coefficient of ASSIGN and SCORE2 was 0.89 (**Supplementary Figure 3**). Given that the ASSIGN score was tailored to Scottish population, we decided to use it as the primary CVD risk measure, with SCORE2 used in sensitivity analyses.

### Incremental model using cardiac troponin and cardiac troponin EpiScores

We tested whether concentrations of cardiac troponin were associated with CVD risk over and above ASSIGN. While measured concentration of cTnI was associated with an HR of 1.20 per SD increase (95% CI 1.13, 1.29; P=1.9×10^-8^), an EpiScore generated for cTnI (see **Supplementary Methods** for details) was weakly associated with the measured concentrations (incremental R^2^=0.10%, P=0.03) and did not predict CVD risk in Cox models adjusted for ASSIGN (P= 0.97). For that reason, it was not considered as a feature in the generation of the composite CVD score.

### Incremental model using EpiScores for plasma protein levels

We then tested whether 109 protein EpiScores generated by Gadd *et al.* (protein description available in **Supplementary Table 2**)^13^ were associated with CVD risk over 16 years of follow-up.

Firstly, we generated 109 Cox PH CVD risk models adjusted for ASSIGN. Each model was additionally adjusted for a different protein EpiScore. Eight EpiScores failed to satisfy the proportional hazards assumption (Schoenfeld residual test P>0.05). Of the remaining 101 protein EpiScores, 67 were significantly associated with CVD risk (P<0.05). After applying a conservative Bonferroni threshold for multiple testing (P<0.05/101 = 5.0×10^-4^), 36 associations remained statistically significant.

Secondly, to understand whether protein EpiScores were associated with CVD risk beyond established biomarkers such as cardiac troponin, we included the concentration of cTnI as a covariate in the model along with ASSIGN and we repeated the analysis. Of the 101 aforementioned protein EpiScores, 65 associated with CVD over and above the ASSIGN score and the concentration of cTnI (P<0.05, **Figure 1**). 33 associations remained significant after correcting for multiple testing.

**Figure 1.**
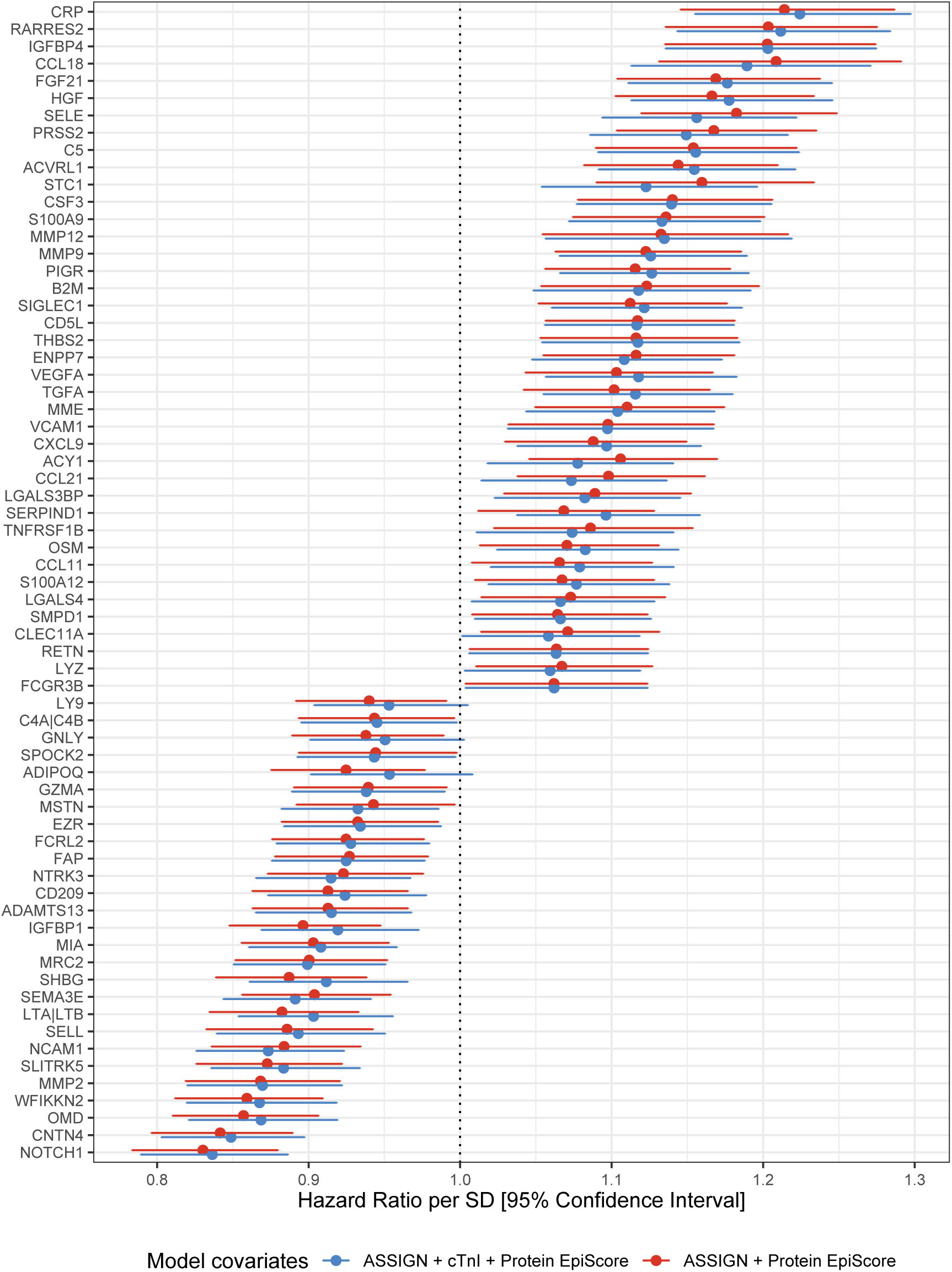
Associations between protein EpiScores and incident CVD. Hazard ratios are plotted for the 67 significant associations (P<0.05) with 95% confidence interval limits. Basic models were adjusted for ASSIGN (blue), whereas full models included the ASSIGN score and concentration of cTnI as covariates (red).

In models adjusted for ASSIGN and the concentration of cTnI, higher levels of 41 protein EpiScores were associated with an increased hazard of CVD (HR>1 and P<0.05). For example, elevated levels of *CRP* and *MMP12* were associated with HR of 1.23 (95% CI 1.16, 1.30; P=9.2×10^-12^) and 1.13 (95% CI 1.06, 1.22, P=5.4×10^-4^) (**Figure 2, A**), respectively. In contrast, higher levels of 24 protein EpiScores were associated with a decreased hazard of CVD (HR<1 and P<0.05). Examples of protein EpiScores belonging to this group include *NOTCH1* (HR per SD of 0.84, 95% CI 0.79, 0.89; P=1.6×10^-9^) and *OMD* (HR per SD of 0.87, 95% CI 0.82, 0.92; P=1.0×10^-6^). The relationships between individual EpiScores and CVD risk have been visualised in a form of risk-over-time (**Figure 2, B**), forest, and Kaplan Meier plots in an online R application: https://shiny.igc.ed.ac.uk/3d2c8245001b4e67875ddf2ee3fcbad2/.

**Figure 2.**
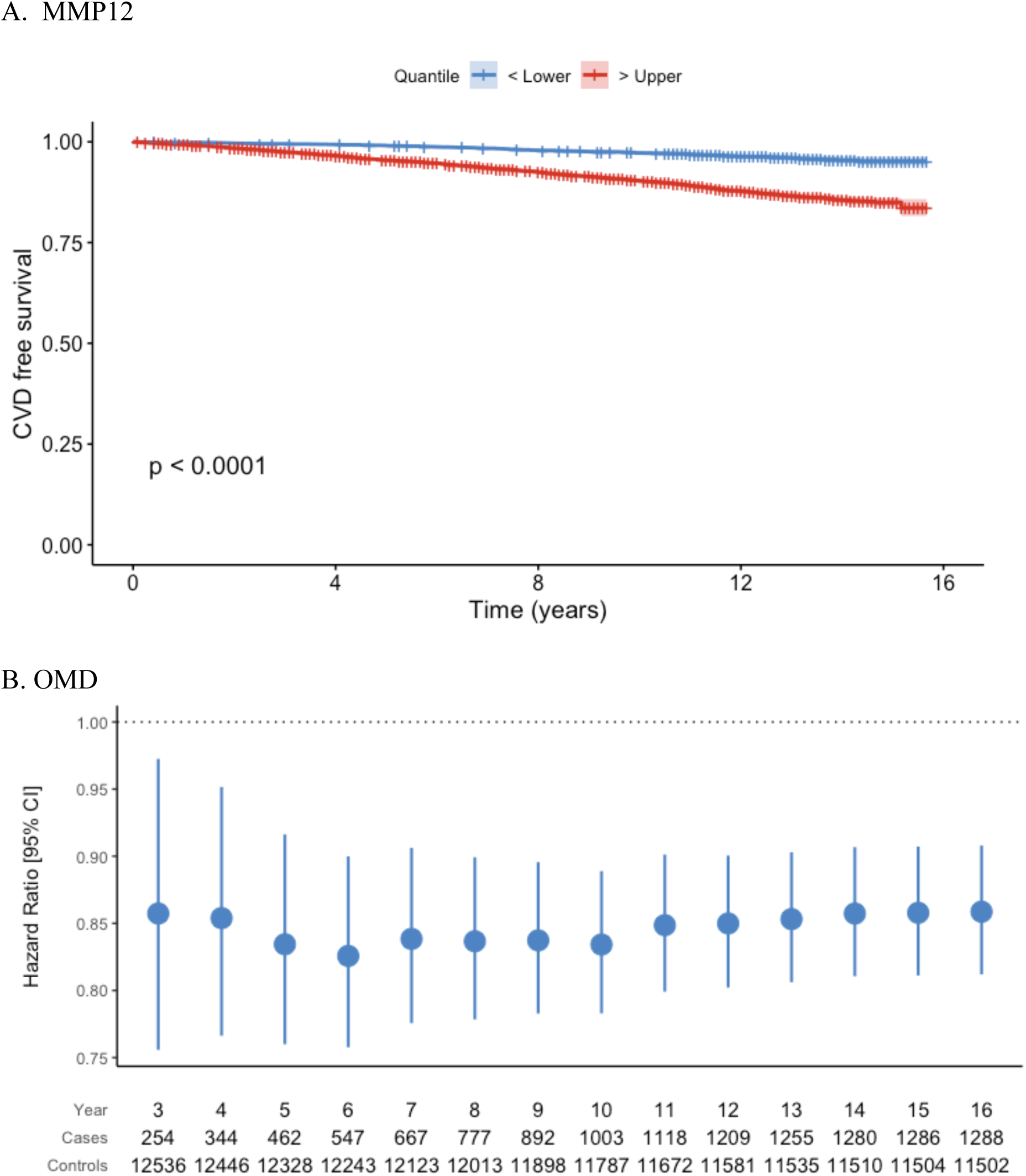
Changes in CVD free survival (A) and CVD risk (B) plotted for two selected protein EpiScores. A. Individuals with higher levels of MMP12 (>75^th^ percentile) had shorter CVD free survival when compared to those with higher level of this EpiScore (<25^th^ percentile). B. Hazard ratios (HR) and 95% confidence intervals associated with the levels of OMD EpiScore plotted over time. At all examined time points the association with CVD risk was significant (P<0.05).

Finally, to learn whether individual protein EpiScore can augment CVD prediction beyond established biomarkers and clinical risk prediction tools, we calculated C-statistics for null and full models. While the null model was adjusted for ASSIGN and the concentration of cTnI (C-stat=0.728), the full model also contained the studied protein EpiScore. **Table 2** lists top 10 associations that result in the greatest improvement of CVD risk prediction.

**Table 2.**
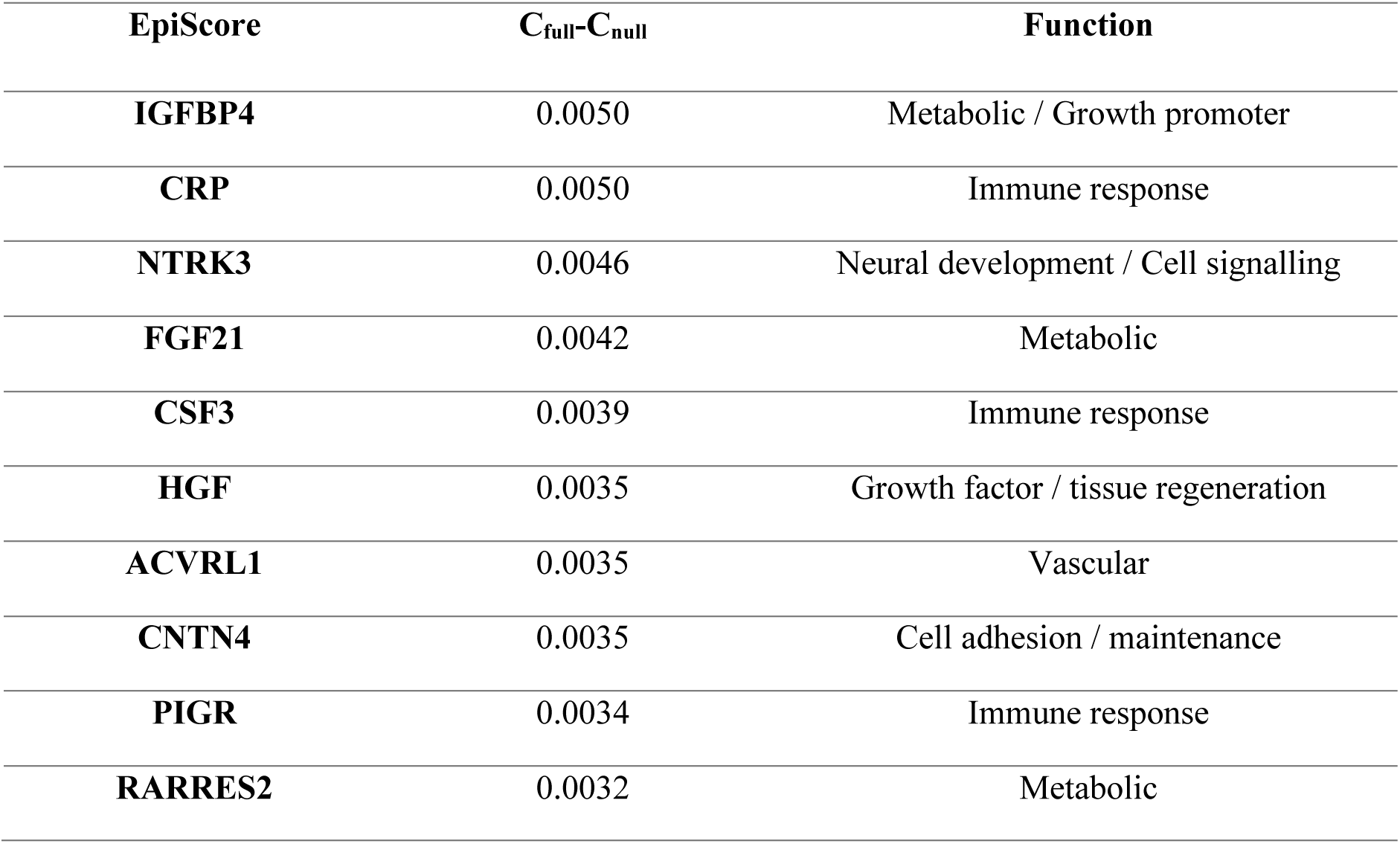
C-statistics calculated for null and full protein EpiScore models. Risk was ascertained over 16 years of follow-up. The null model was adjusted for ASSIGN and the concentration of cardiac troponin I, while the full model also included a studied EpiScore.

| **EpiScore** | **C<sub>full</sub>-C<sub>null</sub>** | **Function** |
| --- | --- | --- |
| **IGFBP4** | 0.0050 | Metabolic / Growth promoter |
| **CRP** | 0.0050 | Immune response |
| **NTRK3** | 0.0046 | Neural development / Cell signalling |
| **FGF21** | 0.0042 | Metabolic |
| **CSF3** | 0.0039 | Immune response |
| **HGF** | 0.0035 | Growth factor / tissue regeneration |
| **ACVRL1** | 0.0035 | Vascular |
| **CNTN4** | 0.0035 | Cell adhesion / maintenance |
| **PIGR** | 0.0034 | Immune response |
| **RARRES2** | 0.0032 | Metabolic |

### Composite Episcore for CVD-risk prediction

To understand whether the above-mentioned protein EpiScores can be used as biomarkers that add additional predictive value over-and-above typically used clinical risk scores (ASSIGN and SCORE2) and the concentration of cTnI, we generated a composite CVD EpiScore – a weighted linear combination of individual protein EpiScores. The score was trained using two modelling techniques: Cox PH Elastic Net and Random Survival Forest. There were 6,880 records in the training set and 3,659 records in the test set. The Elastic Net assigned non-zero coefficients to 45 of 109 protein EpiScores (**Supplementary Table 3**).

In 10-year Elastic Net prediction analysis, the null model (containing age, sex, and ASSIGN) had an AUC of 0.719. The model with the CVD EpiScore increased the AUC to 0.723. The addition of cTnI to the null model resulted in an AUC of 0.721. The full model (null model + cTnI + CVD EpiScore) AUC was 0.724. Full output for the CVD models including C-statistics and a comparison with SCORE2 can be found in **Supplementary Table 4** and **Supplementary Table 5**. The CVD EpiScore remained statistically significant after adjusting for the concentration of cTnI in models incorporating ASSIGN and SCORE2 (HR=1.32, P=3.7×10^-3^ and HR=1.36, P=1.4×10^-3^, respectively).

Random Survival Forest–based analysis (see **Methods**) yielded similar results. The null model (as above) had an AUC of 0.719. Adding the CVD EpiScore to the null model increased the AUC to 0.721. The full model adjusted for CVD EpiScore and the concentration of cardiac troponin had an AUC of 0.723.

## Discussion

In this study, we describe 65 novel epigenetic biomarkers that associate with long-term risk of CVD independently of a clinical risk prediction tool (ASSIGN) and the concentration of an established protein biomarker (cTnI). The most statistically significant EpiScores reflected concentrations of proteins involved in metabolic, immune, and developmental pathways. A weighted linear combination of protein EpiScores (the composite protein CVD EpiScore) was significantly associated with CVD risk in models adjusted for ASSIGN. Although the score may be a useful addition to other omic features in future CVD risk prediction tools, at present it is unlikely to be measured in a clinical setting ^26^.

One previous study focused on how DNAm biomarkers improve CVD risk prediction ^27^. Using time-to-event data and a panel of 60 blood DNAm biomarkers measured in an Italian cohort of 1803 individuals (295 cases), Cappozzo *et al.* trained a composite score for predicting short-term risk of CVD. In comparison, we focused on a more extensive panel of DNAm protein markers in addition to measured troponin levels. We also ran univariate analyses to identify individual proteins and protein classes that associate with CVD. Furthermore, we developed 10-year prediction models (the prediction window for which both ASSIGN and SCORE2 are recommended) trained on more than double the number of cases.

Our findings suggest that individual protein EpiScores capture disease-specific biomarker signals relevant to CVD risk prediction. The relationships found between 65 protein EpiScores and incident CVD mirrored previously reported associations between CVD and measured protein concentrations. For example, elevated levels of *CRP*, a marker for systemic low-grade inflammation, have been associated with multiple age-related morbidities, including CVD ^28^. *MMP12* and *OMD*, in turn, are involved in maintaining the stability of atherosclerotic plaques.

While *MMP12* contributes to the growth and destabilisation of plaques ^29^, increased levels of *OMD* have been observed in macro-calcified plaques from asymptomatic patients ^30^. Finally, multiple studies have demonstrated that *NOTCH1* signalling protects the heart from CVD-induced myocardial damage. The Notch1 pathway is involved in neo-angiogenesis and revascularisation of failing heart ^31^. It limits the extent of ischemic injury ^31^, reduces fibrosis ^32^ and improves cardiac function ^33^. Several of the protein EpiScores associated with CVD in our study, such as *SELE* and *C5*, have also been shown to associate with stroke and ischemic heart disease in our previous work ^13^.

In contrast, the protein EpiScore we trained for cTnI was not associated with the incidence of CVD. Therefore, we excluded it from composite CVD score generation. This highlights an important consideration in the development of multi-omics biomarkers, as there are unlikely to be DNAm differences that associate with every blood protein. For example, the 109 protein EpiScores generated by Gadd *et al* that we make use of in our study were extracted as the best-performing EpiScores from a total set of 953 proteins tested as potential outcomes ^13^. It is therefore not always possible to generate a meaningful protein EpiScore that reflects the protein biology. In the case of cardiac troponins, the elevations in circulating cTnI and cTnT are a result of a leakage of these proteins from the damaged heart muscle into the bloodstream ^34^. As opposed to transcription, this process is not regulated by DNA methylation. Therefore, the methylation signal underlying increased concentration of cardiac troponin in the bloodstream may too weak to enable generation of a meaningful EpiScore.

Strengths of this study include the precise timing of the CVD event through the electronic health records, the ability to generate a clinical risk predictor in a population cohort, and the very large sample size for DNAm, which also permitted the splitting of the data into train/test sets to formally examine the improvement in risk prediction from our omics biomarkers.

Limitations to this work include the generalisability beyond a Scottish population. In this study, we trained and tested predictors in a Scottish cohort to augment the ASSIGN score. However, many of the protein EpiScores were trained in a German cohort (KORA) and projected to Generation Scotland ^13^. Although the ASSIGN score is tailored to the Scottish population, we observed similar findings across all models when replacing it with SCORE2, which is widely used across Europe.

In conclusion, we identified novel epigenetic signals that were associated with the incidence of CVD independently of ASSIGN and the concentration of cardiac troponin. The exploration of associations between protein EpiScores and CVD shed light on the aetiology and molecular biology of the disease. As DNAm and proteins are assessed in increasingly large cohort samples, it will be possible to evaluate more precisely the potential gains in risk prediction, disease prevention and any associated health economic benefits.

## Data Availability

All code used in the analyses is available with open access at the following Gitlab repository: https://github.com/aleksandra-chybowska/troponin_episcores. The source datasets analysed during the current study are not publicly available due to them containing information that could compromise participant consent and confidentiality. Data can be obtained from the data owners. Instructions for accessing Generation Scotland data can be found here: https://www.ed.ac.uk/generation-scotland/for-researchers/access; the 'GS Access Request Form' can be downloaded from this site. Completed request forms must be sent to to be approved by the Generation Scotland access committee. Videos introducing the 109 protein EpiScores and MethylDetectR application are available at https://www.youtube.com/channel/UCxQrFFTIItF25YKfJTXuumQ.

## Acknowledgements

We are grateful to all the families who took part, the general practitioners and the Scottish School of Primary Care for their help in recruiting them, and the whole Generation Scotland team, which includes interviewers, computer and laboratory technicians, clerical workers, research scientists, volunteers, managers, receptionists, healthcare assistants and nurses. We would also like to thank Dr Robert Hillary for helpful feedback on the manuscript and analyses.

## Funding

This research was funded in whole, or in part, by the Wellcome Trust (104036/Z/14/Z, 108890/Z/15/Z, 220857/Z/20/Z, and 216767/Z/19/Z). For the purpose of open access, the author has applied a CC BY public copyright licence to any Author Accepted Manuscript version arising from this submission. A.D.C is supported by a Medical Research Council PhD Studentship in Precision Medicine with funding by the Medical Research Council Doctoral Training Programme and the University of Edinburgh College of Medicine and Veterinary Medicine.

D.A.G. is funded by the Wellcome Trust 4-year PhD in Translational Neuroscience–training the next generation of basic neuroscientists to embrace clinical research (108890/Z/15/Z).

E.B. and R.E.M. are supported by the Alzheimer’s Society major project grant AS-PG-19b-010.

Generation Scotland received core support from the Chief Scientist Office of the Scottish Government Health Directorates (CZD/16/6) and the Scottish Funding Council (HR03006). Genotyping and DNA methylation profiling of the GS samples was carried out by the Genetics Core Laboratory at the Clinical Research Facility, University of Edinburgh, Scotland and was funded by the Medical Research Council UK and the Wellcome Trust (Reference 104036/Z/14/Z, 220857/Z/20/Z, and 216767/Z/19/Z). The DNA methylation profiling and analysis was supported by Wellcome Investigator Award 220857/Z/20/Z and Grant 104036/Z/14/Z (PI: AM McIntosh) and through funding from NARSAD (Ref: 27404; awardee: Dr DM Howard) and the Royal College of Physicians of Edinburgh (Sim Fellowship; Awardee: Dr HC Whalley). All components of GS:SFHS, including the protocol and written study materials, have received formal, national ethical approval from NHS Tayside Research Ethics Committee (REC reference number 05/S1401/89). In addition local approval has been obtained from NHS Glasgow Research Ethics Committee, and from NHS Glasgow and NHS Tayside Research and Development Offices, as is required.

## Author contributions

R.E.M., and A.D.C. were responsible for the conception and design of the study. A.D.C carried out the data analyses. D.A.G, Y.C and E.B. contributed to the analyses and methodology.

R.E.M. and A.D.C. drafted the article. A.C. facilitated data linkage. K.L.E. and J.F.P. were involved in conceptualisation and provided consultation on the methodology. S.W.M., R.M.W., N.W., L.M., C.S., A.W.M., K.L.E contributed to data collection and preparation. All authors read and approved the final manuscript.

## Conflict of interest

R.E.M. and L.M. have received a speaker fee from Illumina. R.E.M. is an advisor to the Epigenetic Clock Development Foundation. D.A.G. and R.E.M. have received consultancy fees from Optima partners. All other authors declare no competing interests.

## Supplement

**Supplementary Figure 1.**
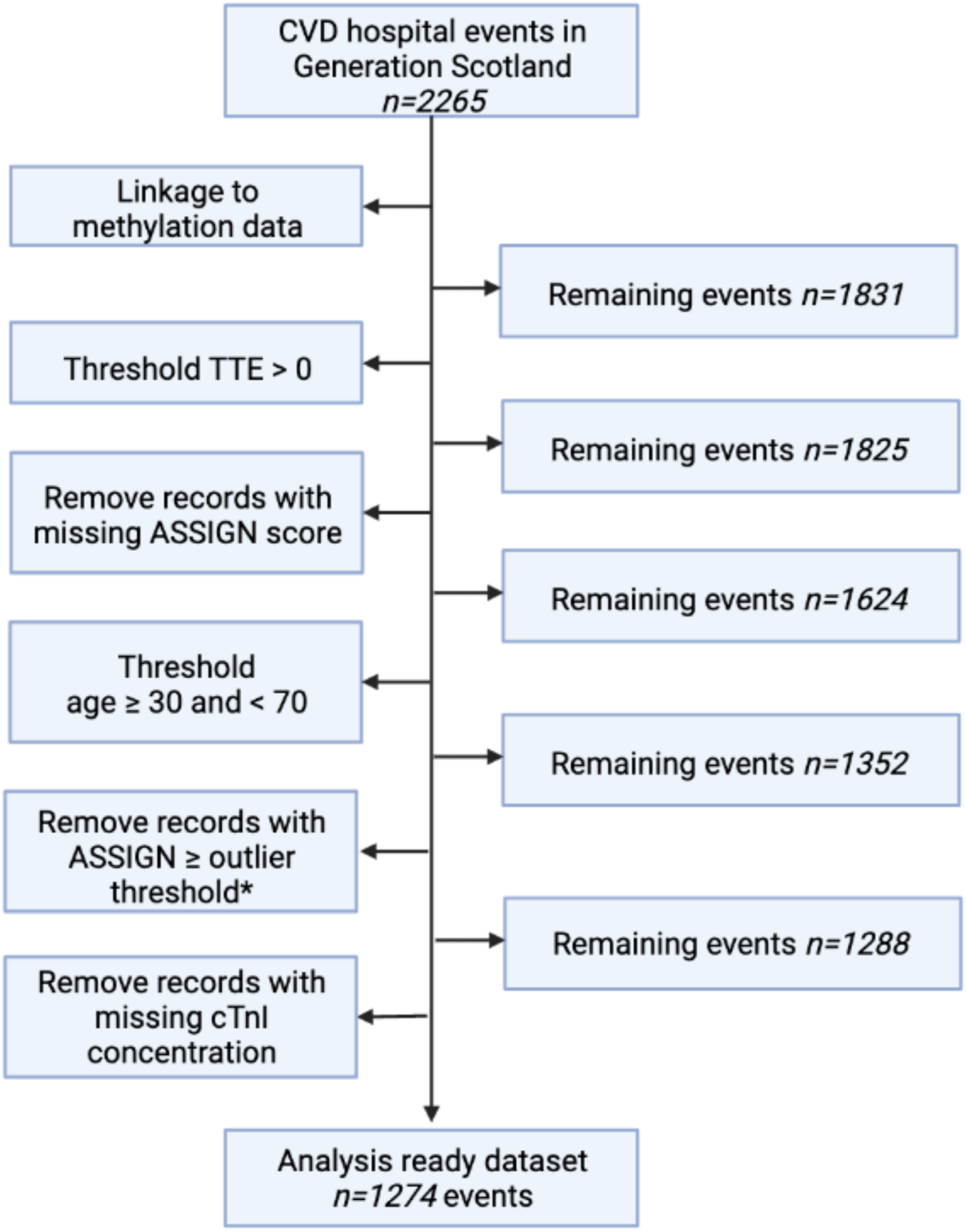
Pre-processing of the full dataset. This pipeline was used to prepare data for Cox PH models aimed at the identification of potential predictors of CVD. *Outlier threshold – an ASSIGN value that is more than 3 SD from the mean. Created with BioRender.com.

**Supplementary Figure 2.**
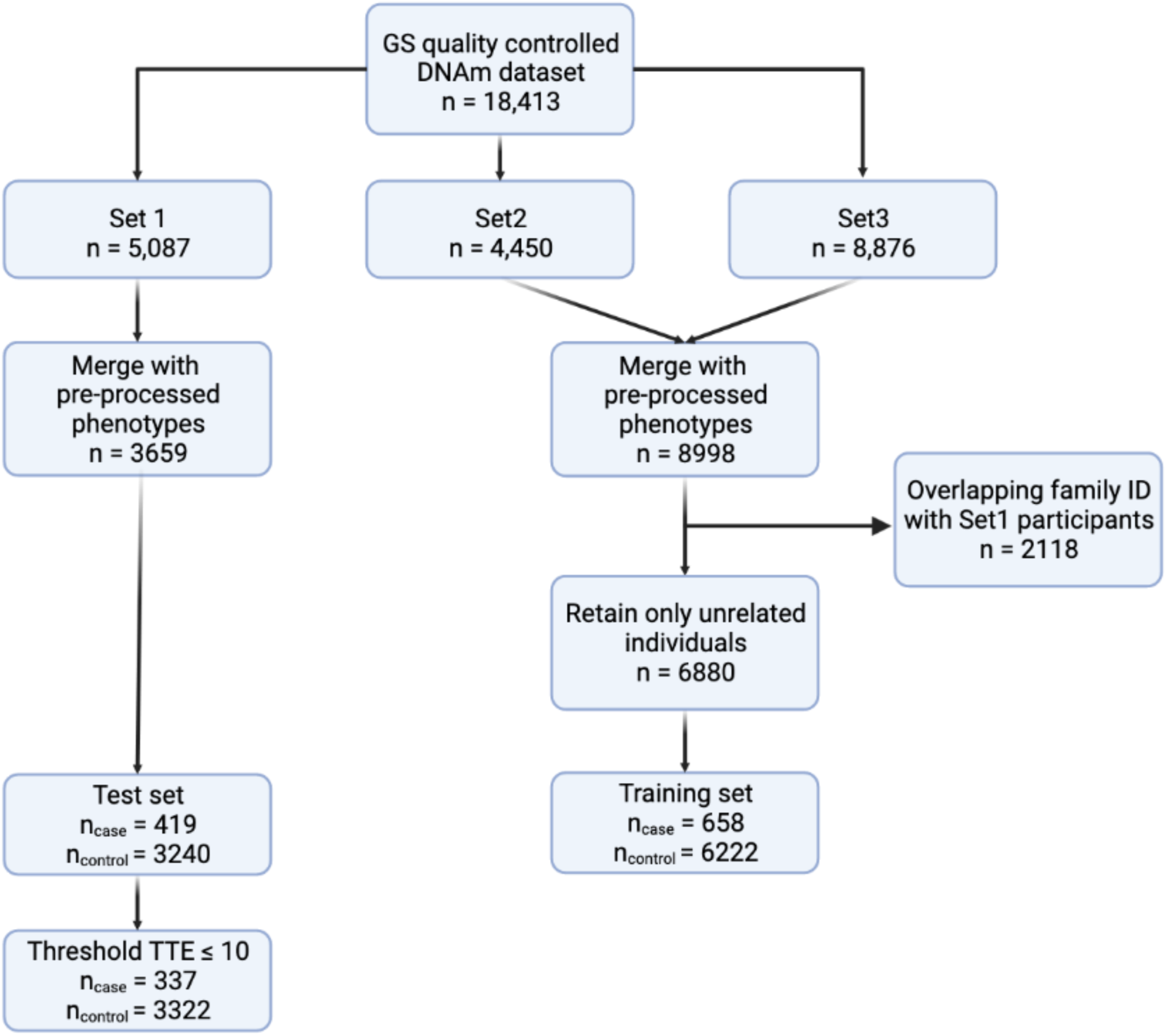
Pre-processing of the training and test sets. While Set 1 was the test set, the training set consisted of Sets 2 and 3 combined. Pre-processing phenotypes (ASSIGN components, composite CVD event status and time-to-event) consisted of filtering out records with time-to-event≤0, removing records with a missing ASSIGN score, thresholding age of participants to 30-70 years and removing outliers (ASSIGN value that is more than 3 SD from the mean). To ensure that there were no individuals with overlapping family ids across the training and the test set, any individuals in the training set associated with the same family id as individuals from the test set were excluded from further analyses. Created with BioRender.com.

**Supplementary Figure 3.**
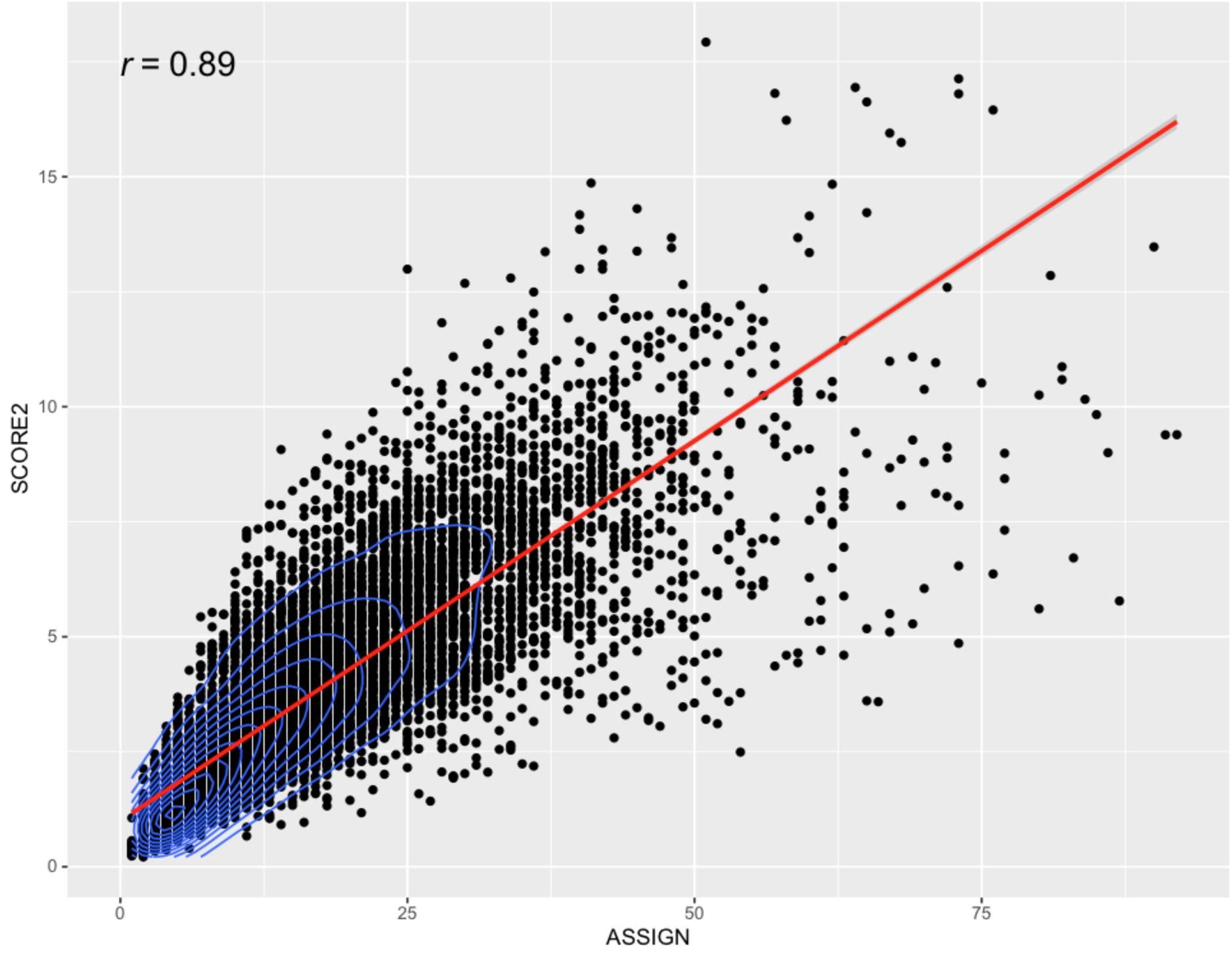
Strongly correlated 10-year CVD risk estimates produced by ASSIGN and SCORE2.

**Supplementary Table 1.**
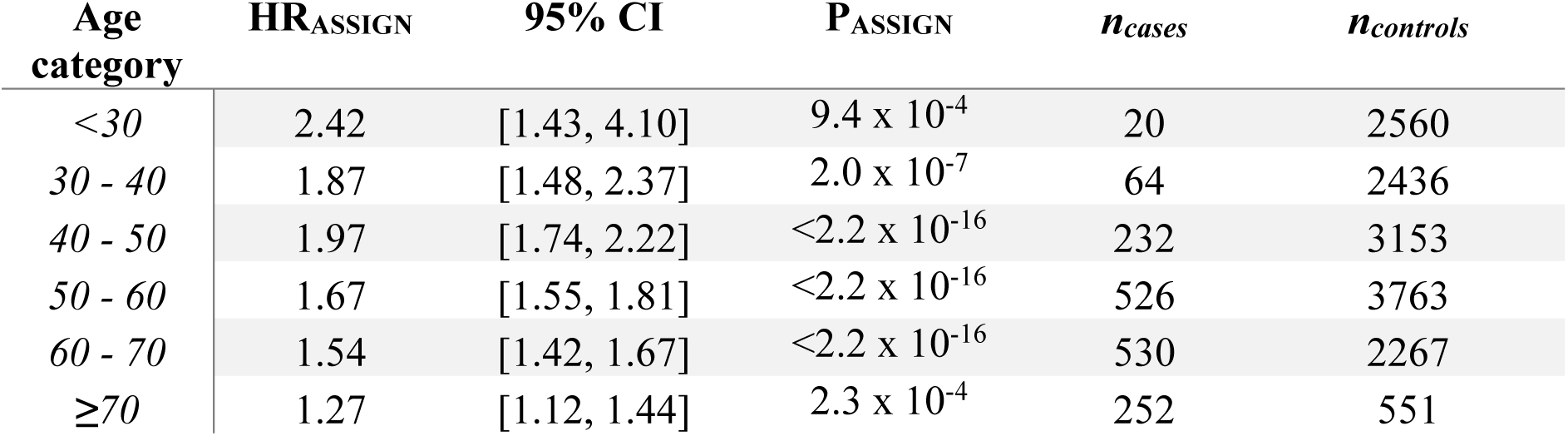
Hazard ratio (HR) of ASSIGN estimated by Cox proportional hazards CVD model adjusted for relatedness using kinship matrix where the ASSIGN score is the only predictor.

| **Age category** | **HR<sub>ASSIGN</sub>** | **95% CI** | **P<sub>ASSIGN</sub>** | ***n*<sub>cases</sub>** | ***n*<sub>controls</sub>** |
| --- | --- | --- | --- | --- | --- |
| <30 | 2.42 | [1.43, 4.10] | 9.4 x 10<sup>-4</sup> | 20 | 2560 |
| 30 - 40 | 1.87 | [1.48, 2.37] | 2.0 x 10<sup>-7</sup> | 64 | 2436 |
| 40 - 50 | 1.97 | [1.74, 2.22] | <2.2 x 10<sup>-16</sup> | 232 | 3153 |
| 50 - 60 | 1.67 | [1.55, 1.81] | <2.2 x 10<sup>-16</sup> | 526 | 3763 |
| 60 - 70 | 1.54 | [1.42, 1.67] | <2.2 x 10<sup>-16</sup> | 530 | 2267 |
| ≥70 | 1.27 | [1.12, 1.44] | 2.3 x 10<sup>-4</sup> | 252 | 551 |

**Supplementary Table 2.**
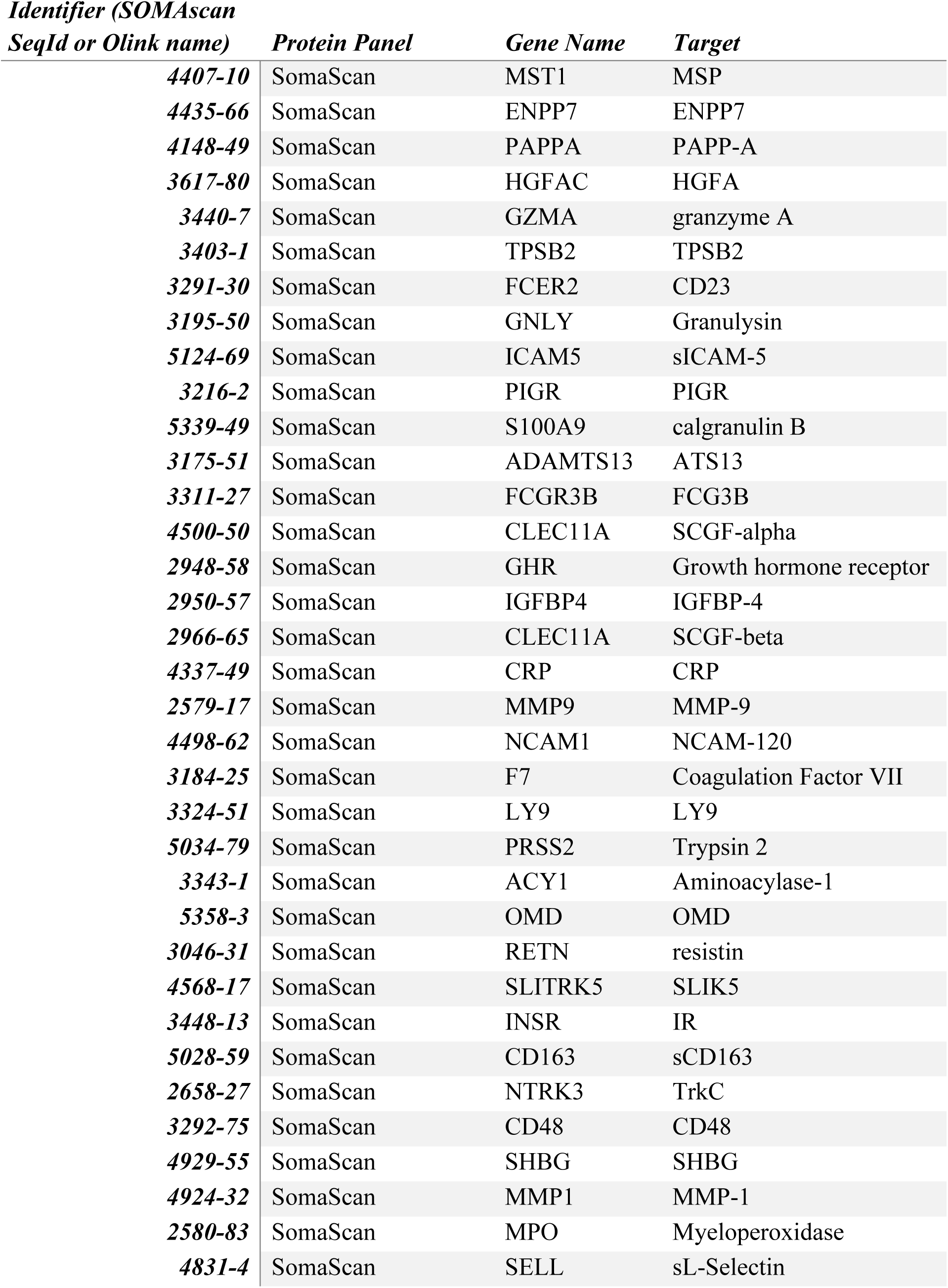
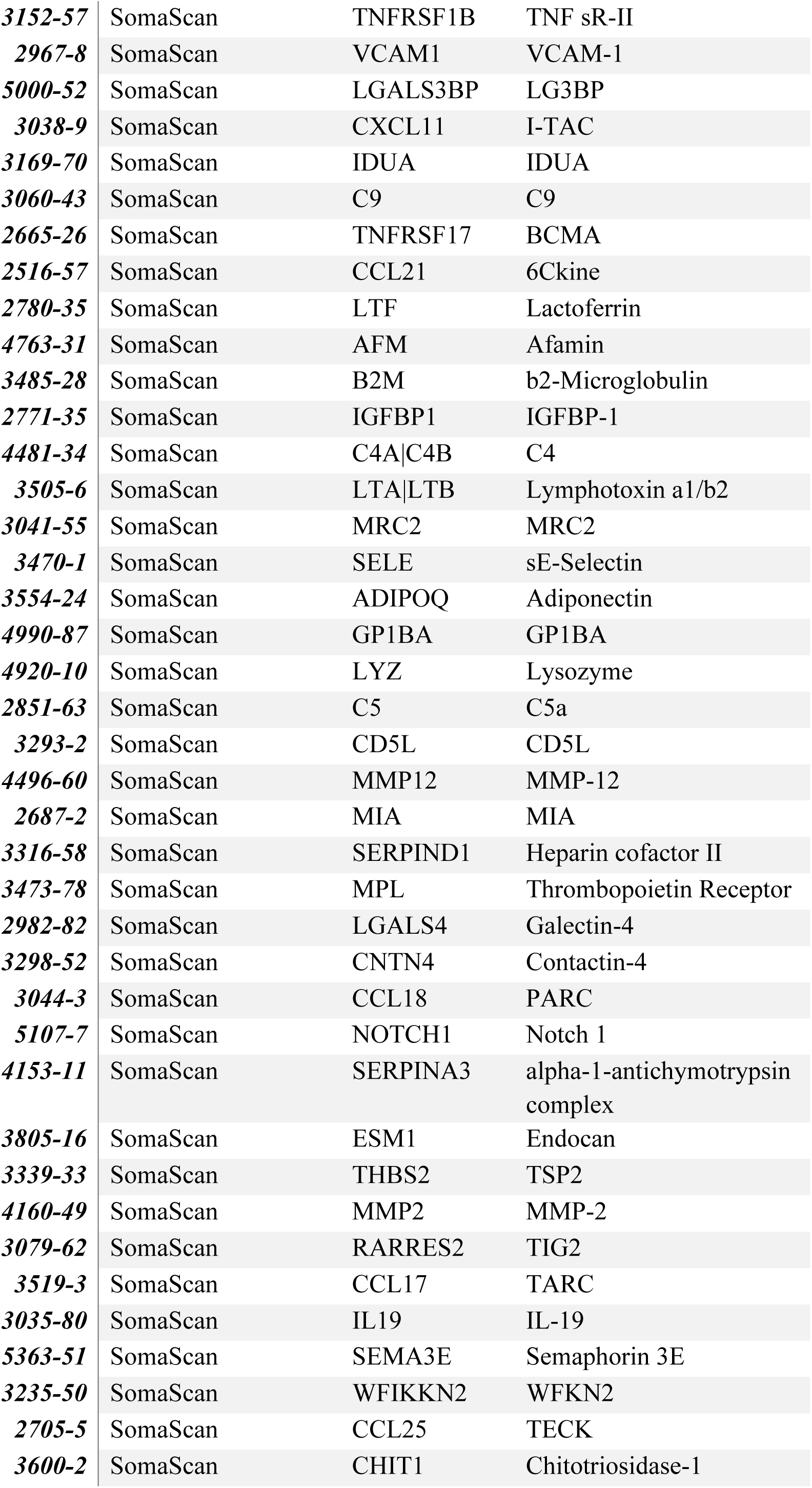
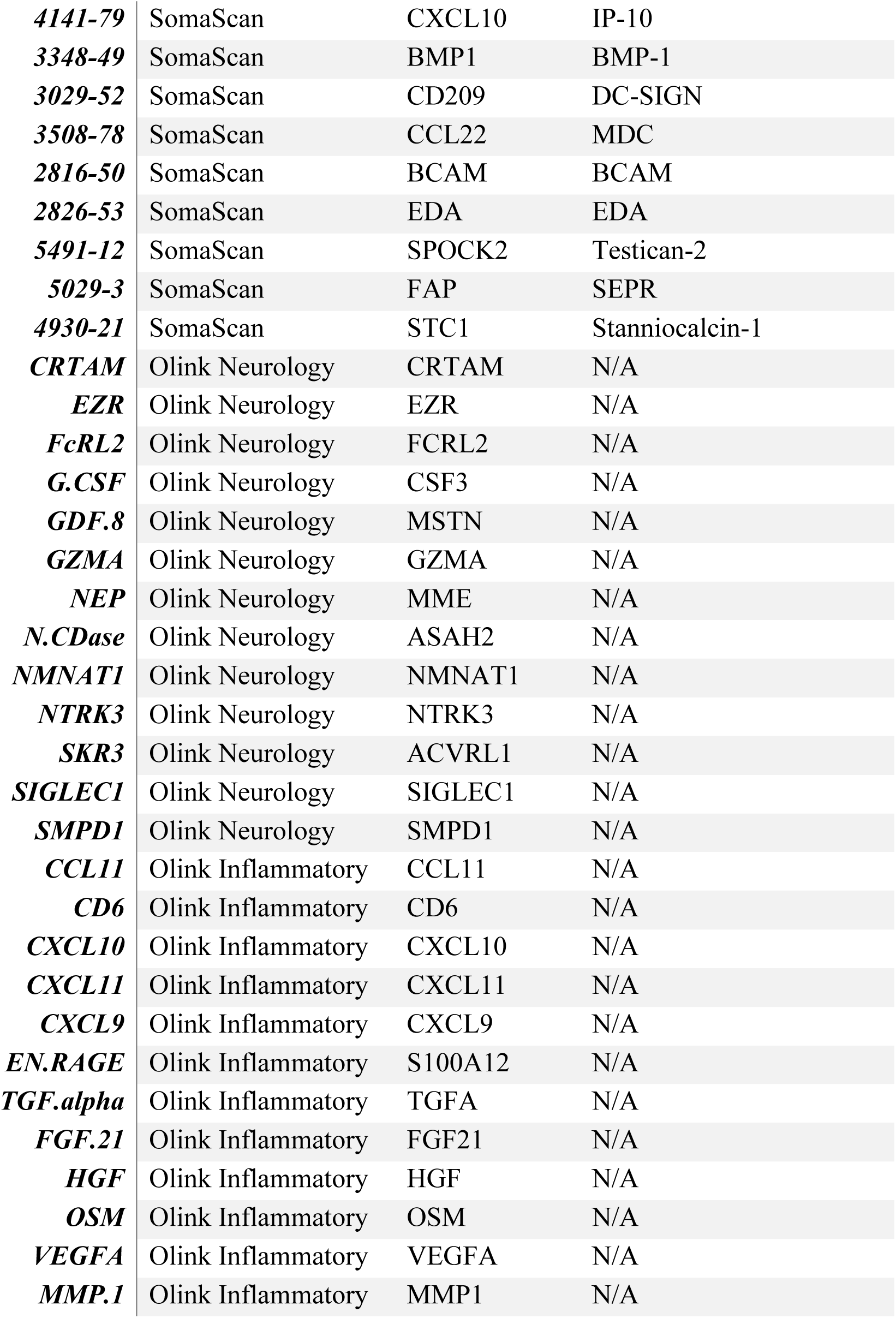
Description of the 109 proteins for which Gadd et al. generated robust EpiScores. Table adapted from Gadd *et. al* ^13^.

**Supplementary Table 3.** Predictor weights for the 45 protein EpiScores included in the.

| *Variable* | *Coefficient* |
| --- | --- |
| *GZMA* | -0.1070275 |
| *N.CDase* | -0.0013707 |
| *NTRK3* | -0.0105284 |
| *SKR3* | 0.02292682 |
| *CCL11* | 0.01486593 |
| *CXCL11* | -0.0363625 |
| *FGF.21* | 0.05884652 |
| *HGF* | 0.05611636 |
| *MMP.1* | -0.0528479 |
| *2705-5* | 0.0411362 |
| *2851-63* | 0.07807877 |
| *2948-58* | 0.03807306 |
| *2967-8* | 0.08372915 |
| *3029-52* | -0.0004437 |
| *3038-9* | -0.0062101 |
| *3044-3* | 0.2568256 |
| *3060-43* | -0.078839 |
| *3079-62* | 0.1024936 |
| *3152-57* | 0.01995523 |
| *3175-51* | 0.02976706 |
| *3291-30* | 0.03575731 |
| *3292-75* | -0.0045541 |
| *3293-2* | 0.04389995 |
| *3339-33* | 0.04065941 |
| *3348-49* | 0.02393257 |
| *3403-1* | -0.0070861 |
| *3448-13* | 0.02739967 |
| *3470-1* | 0.10485373 |
| *3485-28* | 0.05615907 |
| *3505-6* | -0.0178471 |
| *3519-3* | -0.0543479 |
| *3600-2* | 0.03783531 |
| *3617-80* | 0.03982561 |
| *4160-49* | -0.0499596 |
| *4407-10* | 0.03860081 |
| *4435-66* | 0.01144093 |
| *4481-34* | -0.0549405 |
| *4496-60* | 0.10710256 |
| *4763-31* | 0.03997106 |
| *4831-4* | -0.0265954 |
| *4920-10* | -8.29E-05 |
| *4924-32* | 0.0123399 |
| *5000-52* | 0.00608975 |
| *5124-69* | 0.00134476 |
| *5363-51* | -0.0156491 |

**Supplementary Table 4.** Summary of CVD risk models based on ASSIGN. N – null model (adjusted for age + sex + ASSIGN), E – CVD EpiScore, T – cardiac troponin I.

| **Model** | **HR<sub>age</sub>** | **P<sub>age</sub>** | **HR<sub>sex</sub>** | **P<sub>sex</sub>** | **HR<sub>ASSIGN</sub>** | **P<sub>ASSIGN</sub>** | **HR<sub>E</sub>** | **P<sub>E</sub>** | **HR<sub>T</sub>** | **P<sub>T</sub>** | **AUC** | **PRAUC** | **C-index** |
| --- | --- | --- | --- | --- | --- | --- | --- | --- | --- | --- | --- | --- | --- |
| **N** | 1.01 | 0.54 | 1.13 | 0.23 | 2.05 | <2.2x10<sup>-16</sup> | - | - | - | - | 0.719 | 0.198 | 0.712 |
| **N+E** | 1.00 | 0.99 | 1.05 | 0.64 | 1.94 | 1.10x10<sup>-16</sup> | 1.3 | 5.6x10<sup>-3</sup> | - | - | 0.723 | 0.201 | 0.715 |
| **N+T** | 1.00 | 0.83 | 1.04 | 0.71 | 2.02 | <2.2x10<sup>-16</sup> | - | - | 1.16 | 1.1x10<sup>-2</sup> | 0.721 | 0.202 | 0.714 |
| **N+E+T** | 1.00 | 0.65 | 0.96 | 0.71 | 1.91 | 6.70x10<sup>-16</sup> | 1.32 | 3.7x10<sup>-3</sup> | 1.17 | 7.1x10<sup>-3</sup> | 0.724 | 0.205 | 0.717 |

**Supplementary Table 5.** Summary of CVD risk models based on SCORE2. N – null model (adjusted for age + sex + ASSIGN), E – CVD EpiScore, T – cardiac troponin I.

| **Model** | **HR<sub>age</sub>** | **P<sub>age</sub>** | **HR<sub>sex</sub>** | **P<sub>sex</sub>** | **HR<sub>SCORE2</sub>** | **P<sub>SCORE2</sub>** | **HR<sub>E</sub>** | **P<sub>E</sub>** | **HR<sub>T</sub>** | **P<sub>T</sub>** | **AUC** | **PRAUC** | **C-index** |
| --- | --- | --- | --- | --- | --- | --- | --- | --- | --- | --- | --- | --- | --- |
| **N** | 1.02 | 0.05 | 0.96 | 0.75 | 1.79 | 1.8x10<sup>-11</sup> | - | - | - | - | 0.699 | 0.174 | 0.696 |
| **N+E** | 1.01 | 0.15 | 0.92 | 0.50 | 1.64 | 5.4x10<sup>-8</sup> | 1.34 | 2.4x10<sup>-3</sup> | - | - | 0.704 | 0.179 | 0.699 |
| **N+T** | 1.01 | 0.14 | 0.88 | 0.32 | 1.77 | 6.3x10<sup>-11</sup> | - | - | 1.18 | 4.8x10<sup>-3</sup> | 0.701 | 0.175 | 0.698 |
| **N+E+T** | 1.01 | 0.37 | 0.84 | 0.16 | 1.61 | 1.8x10<sup>-7</sup> | 1.36 | 1.4 x10<sup>-3</sup> | 1.19 | 2.7 x10<sup>-3</sup> | 0.707 | 0.182 | 0.702 |

## Supplementary Methods

### EpiScores for cardiac troponin

We generated an EpiScore for cTnI. The training set consisted of Sets 2 and 3 combined, whereas Set 1 was the test set. To minimise information from shared environments and relatedness leaking between the training and test sets, any individuals in the training set associated with the same family ID as individuals from the test set were excluded from further analyses. After this step, there were 9,754 individuals in the training set and 5,003 individuals in the test set.

As part of data pre-processing, all variables were trimmed of outliers (points beyond 3 SDs of the mean). The measured concentrations of cTnI underwent rank-based inverse normal transformation. Using linear mixed effects models, age, age^2^ and sex were regressed out of the transformed variable, with a pedigree-based kinship matrix fitted as a random effect. The subsequent model residuals were saved and entered as the outcome of elastic net regression models. Methylation levels were treated as the independent variables. Elastic net models were run using biglasso (version 1.5.0) in R (version 4.0.3). The L1-L2 mixing parameter was set to alpha=0.5, and ten-fold cross-validation was applied. Non-zero coefficients/weights from the models were extracted. To calculate the EpiScore, CpG methylation values were multiplied by these coefficients and summed for each individual in the test set. The proportion of variance explained by the EpiScore was estimated by comparing R^2^ estimates from the null and full linear regression models. Null models were adjusted for age, sex, and cTnI whereas full models accounted for age, sex, cTnI and the EpiScore.

### EpiScores for 109 circulating proteins

We have previously generated 109 EpiScores for plasma protein levels ^13^. Briefly, elastic net penalised regression models were run with 953 possible protein levels as outcomes and up to 428,489 DNAm measurements from the Illumina 450k array as input features. Protein levels were adjusted for genetic effects (protein quantitative trait loci – pQTLs) prior to training EpiScores. Protein EpiScores were trained in two cohorts: the Lothian Birth Cohort of 1936 (training set contained between 725 and 875 individuals for 160 Olink inflammatory and neurology proteins) and the German cohort, KORA (944 individuals, 793 SOMAscan proteins). 109 scores (84 trained in KORA and 25 trained in the Lothian Birth Cohort 1936) explained between 1% and 58% of the variance in protein levels (with r>0.1 and P<0.05) in independent test cohorts. In this study, the 109 protein EpiScores were projected into Generation Scotland (n = 18,413) through the publicly available MethylDetectR Shiny App. Before applying weights, DNAm level at each site was scaled to have a unit SD and mean of zero. This procedure was performed separately in each set.

## Notes

### Author Declarations

All components of GS:SFHS, including the protocol and written study materials, have received formal, national ethical approval from NHS Tayside Research Ethics Committee (REC reference number 05/S1401/89). In addition local approval has been obtained from NHS Glasgow Research Ethics Committee, and from NHS Glasgow and NHS Tayside Research and Development Offices, as is required.

### Summary of Updates

- Results section: EpiScore models further adjusted for the concentration of cardiac troponin I - Figure 1 revised to include models corrected for ASSIGN and the concentration of cardiac troponin I - Section on EpiScores associated with CVD expanded

